# Undocumented migrants in the Netherlands: adverse life events, living conditions and mental health

**DOI:** 10.1101/2020.07.16.20148759

**Authors:** Sandrine JC Vollebregt, Willem F Scholte, Annette Hoogerbrugge, Koen Bolhuis, Jentien Vermeulen

## Abstract

**Background:** Undocumented migrants have worse health and living conditions and poor access to health care.

**Aims and objectives:** To examine the prevalence and determinants of mental health problems in undocumented migrants.

**Design:** Observational study, integrating cross-sectional questionnaire data with retrospective electronic patient record data.

**Methods:** Undocumented migrants attending medical and psychological consultation hours of a Netherlands-based non-governmental organization, completed the Self-Reporting Questionnaire (SRQ). We examined scores of the instrument’s 24 items version (SRQ-24) and its 20 items version (SRQ-20). Correlations with determinants were estimated using parametric tests.

**Results:** On the SRQ-20, 85% of the sample (N=101) scored ≥8, the clinical cut-off value; mean=12.4 ± 4.6, range 0-20. Adverse life events like physical and sexual assault were reported in 37% of the medical records (N=99) and had a medium to large effect (Cohen’s d=0.76) on SRQ-24 scores.

**Discussion:** Mental health problems are common in undocumented migrants. Adverse life-events are critical determinants.

## Background

Approximately 35.530 undocumented migrants (UMs) resided in the Netherlands according to the latest, but relatively dated, estimate of 2012-2013.(1) Lives of UMs are characterized by unstable living conditions, fear of being arrested, low levels of emotional support and poorer (mental) health.(2–5) Data on mental health is vital to assess health care needs. However, quantitative data on mental health of UMs is scarce and often biased by selection and attrition, as UMs are notoriously difficult to include in regular clinical research. While targeting UMs visiting a low-threshold non-governmental organization’s health care program, this study describes an otherwise poorly visible population. The aims of this study were to examine the prevalence of mental health problems in UMs in the Netherlands, and to identify possible determinants. It was hypothesized that mental health problems were common and related to poorer living conditions and adverse life events.

## Theoretical framework

UMs are a heterogenous group, which comprises rejected asylum seekers, individuals who have not (yet) submitted an application for asylum and overstayers of visas or work permits.(6) The legal position of UMs leads to a precarious existence, as is displayed by a multi-country survey study carried out through Europe.(5) Living and working legally is impossible, which makes housing and working conditions unstable and tough. Almost half of the population has no income from work. UMs are often isolated from the host population, and experience low levels of emotional support.(5) A scoping review shows that UMs report poorer health, and in particular mental health problems are common.(2) UMs consider mental health problems as a direct result of their poor living conditions and illegal status.(7,8) Dutch health care professionals confirm that stress, traumas, sleeping problems and gloominess are more common than in the general population.(3,9)

There is ample evidence on the prevalence of potentially traumatic experiences in refugees.(10,11) A study among rejected Iraqi asylum seekers in Denmark showed an average of 8.5 traumatic events before arrival in the host country.(12) A large systematic review assessing forced migrants yielded prevalence rates of torture and other war-related traumatic events ranging from 1 - 76% (median 27%).(13) It has been shown that such events often have negative mental health consequences, e.g., PTSD, depressions and anxiety disorders.(14,15) Such outcomes may similarly apply for UMs, a substantial proportion of whom have escaped violence and repression.

Despite this poor mental health, mental health care remains difficult to access for UMs in Europe(16), and this seems to be no different in the Netherlands.(7,9) Barriers include insufficient knowledge of and trust in GPs’ competencies regarding mental health, financial and practical barriers, taboo on mental health problems and fear of prosecution.(7,9) In addition, UMs often consider mental health care unlikely to be beneficial in their situation and therefore see it as a last resort, after seeking support from friends or in religion.(7,8)

## Methods

### Participants

Respondents were recruited at medical and at psychological consultation hours of Doctors of the World, a non-governmental organization focussing on health care for UMs. These consultation hours are held in a converted minivan, which visits places in Amsterdam and Rotterdam (two major cities in the Netherlands) where UMs reside, like night shelters, squats and certain neighbourhoods. A convenience sampling method was used. Recruitment and data collection were performed by volunteering medical doctors and psychologists.

The first group, hereinafter referred to as the ‘general group’, consists of people included at the medical consultation hours, before the psychological consultation program came into existence (n=47). They were included regardless of their complaint (whether physical or psychological). The second group, hereinafter referred to as the ‘psychological help-seeking group’ consists of people included after the start of the psychological consultation program. They were included at the medical consultation hours because of presenting a psychological problem (n=16) and at the psychological consultation hours (n=38), resulting in a total of 54 respondents.

### Data Collection

After verbal informed consent was obtained, the Self-Reporting Questionnaire-24 (SRQ-24) was administered. Questions were read aloud in English, Dutch or French. When needed, additional explanation was given.

The first author collected survey data and extracted descriptive data, medical information (presence of chronic disease, psychiatric history, substance use and referral to mental health services) and data about adverse life events from electronic patient files. Patients were included from June 2016 to September 2018. The psychological consultation program was initiated in June 2017.

### Measures

Mental health problems were measured using the Self Reporting Questionnaire (SRQ), a screening instrument for common mental disorders developed by the World Health Organization for the low and middle-income primary health care setting.(17) SRQ items are scored 0 (‘no’, symptom absent) or 1 (‘yes’, symptom present). A sum score is created by the total number of ‘yes’ answers. Higher scores indicate a higher likelihood of a common mental disorder.(17) The original SRQ comprises 25 items, twenty of which are related to neurotic symptoms, four to psychotic symptoms and one to convulsions. Most studies use the 20 items version (SRQ-20), which consists of the neurotic items only. These reflect depressive symptoms, anxiety, and psychosomatic complaints, which, taken together as a group, are often referred to as ‘common mental disorders’. The SRQ-20 has been used and validated in different populations, amongst others in refugee camps.(18–21) We used the SRQ-24 because of the presumed relationship between psychological trauma and psychotic symptoms.(22) For the SRQ-20, usually a cut-off value of 7-8 is used (17); however validation studies in different populations have led to contrasting cut-off values, ranging from ≥3 in Ethiopia to ≥17 for women in Afghanistan.(19,23) We tried to address this limitation by using both the most used cut-off value of ≥8 and a more stringent value of ≥13; the latter is similar to the cut-off value used in a study amongst Afghan mothers caring for children in a refugee camp in Pakistan.(17,21). We found no evidence on cut-off values for the SRQ-24. To enable comparison with other studies, we present both SRQ-20 and SRQ-24 scores.

All electronic patient records were screened for quotes about adverse life events. These quotes were categorized in accordance with the Life Events Checklist for DSM-5.(24)

### Analysis

First, descriptive statistics were used to describe the population. Distribution assessment was performed for the SRQ-20 and the SRQ-24 by making a histogram, comparing means with medians and performing Kolmogorov-Smirnov tests. As scores were distributed normally, means and standard deviations were calculated. The unpaired two-samples t-test was used to compare SRQ-scores of the general group with those of the psychological help-seeking group. Second, Cronbach’s alpha coefficient was calculated to assess internal consistency of the questionnaire. Third, to assess the association between SRQ-20 scores and separate variables, different parametric tests were used: Pearson’s rho (age and length of stay in the Netherlands), one-way ANOVA (living situation and region of origin) and the unpaired two-samples t-test (chronic disease, psychiatric history, substance use, referral to mental health services and adverse life events in the electronic patient record). Effect sizes were calculated using a Cohen’s d calculator.(25) IBM SPSS Statistics version 25 was used for all other statistical analyses.(26)

### Ethical Procedures

This study has been submitted to the Medical Research Ethics Committee of UMC Utrecht, which granted exemption of the obligation to request official ethical approval, meaning the Medical Research Involving Human Subjects Act (WMO, 1999) did not apply to this study. All procedures performed in this study were in accordance with the ethical standards of the 1964 Helsinki declaration and its later amendments or comparable ethical standards. Respondents were informed about the use of anonymized data for analysis and reporting.

## Results

The convenience sample consisted of 76 males and 25 females. The median age was 36 years (interquartile range=29-48). Most respondents originated from the Middle East and North Africa (33%) and Eastern Africa (29%). Thirty-five different home countries were reported. Somalia (11%), Eritrea (11%), Sudan (8%) and Nigeria (8%) were most common. Most people lived in night shelters (33%) or were lodging with acquaintances, friends or family (21%). Sixteen percent lived in a squat or another unofficial location and 9% lived on the streets. The median duration of residence in the Netherlands was 7.5 years (interquartile range=2.3-19.8), but this was only reported in 24% of the cases (Table 1).

**Table 1.**
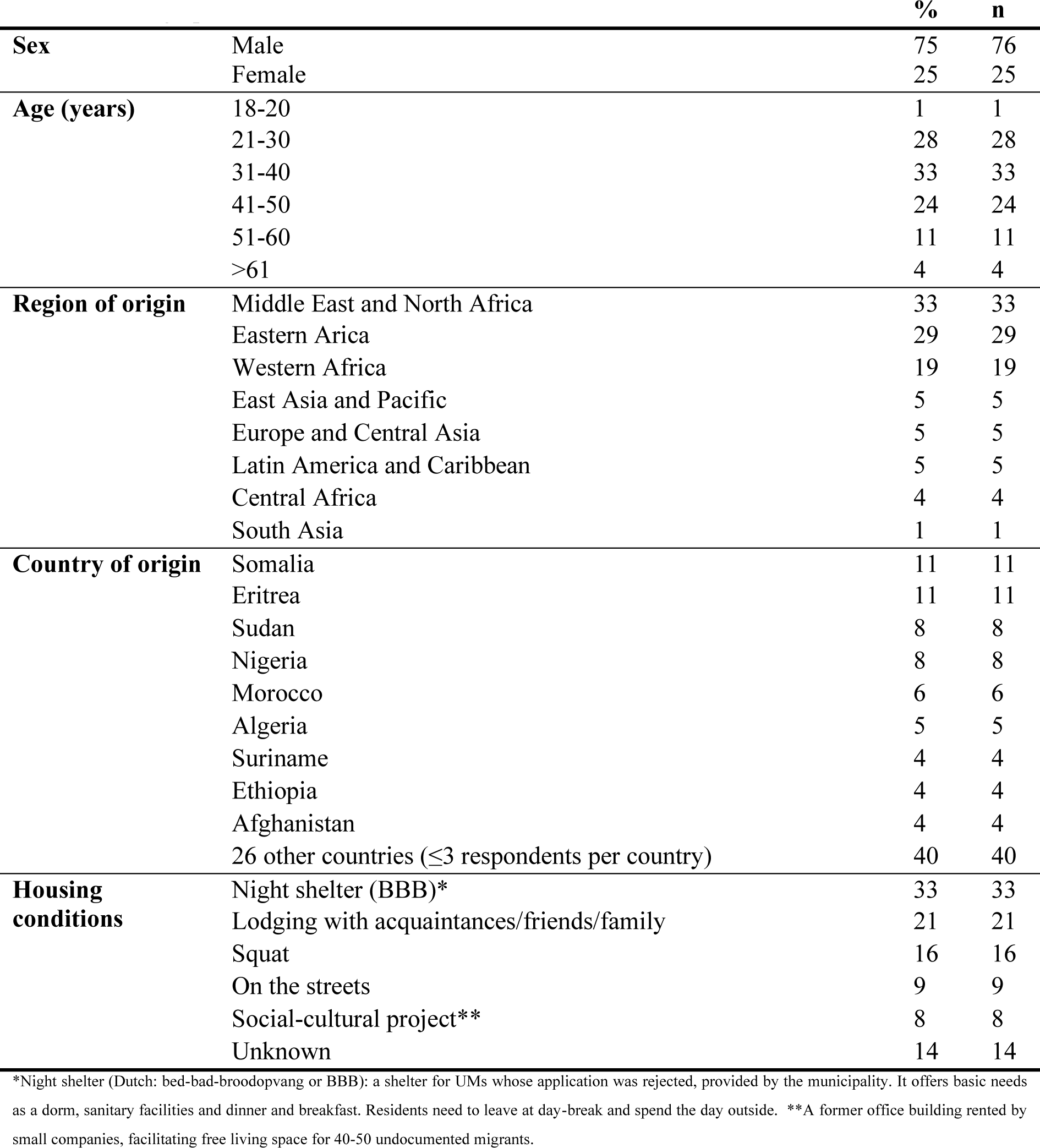
Demographic characteristics (n=101)

| **Table 1** Demographic characteristics (n=101) |  | **%** | **n** |
| --- | --- | --- | --- |
| **Sex** | Male | 75 | 76 |
|  | Female | 25 | 25 |
| **Age (years)** | 18-20 | 1 | 1 |
|  | 21-30 | 28 | 28 |
|  | 31-40 | 33 | 33 |
|  | 41-50 | 24 | 24 |
|  | 51-60 | 11 | 11 |
|  | >61 | 4 | 4 |
| **Region of origin** | Middle East and North Africa | 33 | 33 |
|  | Eastern Arica | 29 | 29 |
|  | Western Africa | 19 | 19 |
|  | East Asia and Pacific | 5 | 5 |
|  | Europe and Central Asia | 5 | 5 |
|  | Latin America and Caribbean | 5 | 5 |
|  | Central Africa | 4 | 4 |
|  | South Asia | 1 | 1 |
| **Country of origin** | Somalia | 11 | 11 |
|  | Eritrea | 11 | 11 |
|  | Sudan | 8 | 8 |
|  | Nigeria | 8 | 8 |
|  | Morocco | 6 | 6 |
|  | Algeria | 5 | 5 |
|  | Suriname | 4 | 4 |
|  | Ethiopia | 4 | 4 |
|  | Afghanistan | 4 | 4 |
| | 26 other countries ($\leq 3$ respondents per country) | 40 | 40 |
| **Housing conditions** | Night shelter (BBB)* | 33 | 33 |
|  | Lodging with acquaintances/friends/family | 21 | 21 |
|  | Squat | 16 | 16 |
|  | On the streets | 9 | 9 |
|  | Social-cultural project** | 8 | 8 |
|  | Unknown | 14 | 14 |

\*Night shelter (Dutch: bed-bad-broodopvang or BBB): a shelter for UMs whose application was rejected, provided by the municipality. It offers basic needs as a dorm, sanitary facilities and dinner and breakfast. Residents need to leave at day-break and spend the day outside. \*\*A former office building rented by small companies, facilitating free living space for 40-50 undocumented migrants.

### Mental Health Problems

The mean SRQ-20 score was 11.6 ± 5.0 in the general group and 13.1 ± 4.3 in the psychological help-seeking group (t(99)=-1.73, p=0.09). The mean SRQ-24 score was 12.4 ± 5.4 in the general group and 14.6 ± 4.8 in the psychological help-seeking group (t(99)=-2.17, p=0.03), indicating that groups differed mostly on the presence of psychotic experiences (Table 2). In the general group, 81% scored above the SRQ-20’s cut-off value of ≥8, versus 87% in the psychological help-seeking group. If the more cautious value of ≥13 is used, 43% of the general group scored above the cut-off value, versus 62% in the psychological help-seeking group (Table 3). The two most reported symptoms were sleeping badly (84%) and feeling nervous, tense or worried (82%). Thirty-seven percent of the respondents had thought about ending their life in the past thirty days. Thirty-five percent reported hearing voices that others could not hear (Table 4).

**Table 2.**
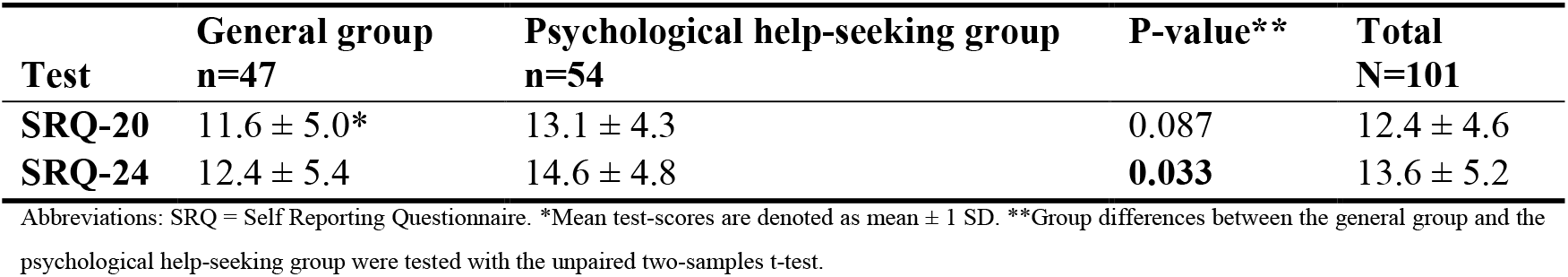
Mean scores SRQ-20 and SRQ-24

| **Test** | **General group**<br>**n=47** | **Psychological help-seeking group**<br>**n=54** | **P-value\*\*** | **Total**<br>**N=101** |
| --- | --- | --- | --- | --- |
| **SRQ-20** | 11.6 ± 5.0* | 13.1 ± 4.3 | 0.087 | 12.4 ± 4.6 |
| **SRQ-24** | 12.4 ± 5.4 | 14.6 ± 4.8 | **0.033** | 13.6 ± 5.2 |

Abbreviations: SRQ = Self Reporting Questionnaire. \*Mean test-scores are denoted as mean ± 1 SD. \*\*Group differences between the general group and the psychological help-seeking group were tested with the unpaired two-samples t-test.

**Table 3.** Percentage of people scoring ≥ cut-off value (SRQ-20)

| **Cut-off value** | **General group**<br>**n=47** | **Psychological help-seeking group**<br>**n=54** | **Total**<br>**n=101** |
| --- | --- | --- | --- |
| **$\geq 8$** | 81% (38)* | 89% (48) | 85% (86) |
| **$\geq 13$** | 43% (20) | 63% (34) | 53% (54) |

Abbreviations: SRQ = Self Reporting Questionnaire. \*Scores are denoted as percentage (n).

**Table 4.** ‘Yes’ answers on SRQ-24*

| **Questions** | **General group**<br>**n=47** | **Psychological help-**<br>**seeking group**<br>**n=54** | **Total group**<br>**n=101** | **Missing**<br>**answers** |
| --- | --- | --- | --- | --- |
| 1. Do you often have headaches? | 78% (37) | 74% (40) | 76% (77) | 0% (0) |
| 2. Is your appetite poor? | 66% (31) | 76% (41) | 71% (72) | 0% (0) |
| 3. Do you sleep badly? | 81% (38) | 89% (47) | 84% (85) | 3% (3) |
| 4. Are you easily frightened? | 67% (30) | 77% (41) | 70% (71) | 1% (1) |
| 5. Do your hands shake? | 28% (13) | 54% (29) | 42% (42) | 1% (1) |
| 6. Do you feel nervous, tense or worried? | 78% (36) | 87% (47) | 82% (83) | 1% (1) |
| 7. Is your digestion poor? | 53% (25) | 56% (29) | 54% (54) | 2% (2) |
| 8. Do you have trouble thinking clearly? | 58% (26) | 82% (44) | 69% (70) | 2% (2) |
| 9. Do you feel unhappy? | 76% (35) | 83% (44) | 78% (79) | 2% (2) |
| 10. Do you cry more than usual? | 45% (21) | 59% (32) | 53% (53) | 0% (0) |
| 11. Do you find it difficult to enjoy your daily activities? | 62% (28) | 72% (36) | 63% (64) | 6% (6) |
| 12. Do you find it difficult to make decisions? | 50% (21) | 60% (32) | 53% (53) | 6% (6) |
| 13. Is your daily work suffering? | 56% (20) | 53% (19) | 39% (39) | 29% (29) |
| 14. Are you unable to play a useful part in life? | 58% (25) | 67% (32) | 56% (57) | 10% (10) |
| 15. Have you lost interest in things? | 53% (24) | 57% (29) | 53% (53) | 5% (5) |
| 16. Do you feel that you are a worthless person? | 56% (24) | 54% (27) | 51% (51) | 8% (8) |
| 17. Has the thought of ending your life been on your mind? | 30% (13) | 45% (24) | 37% (37) | 4% (4) |
| 18. Do you feel tired all the time? | 74% (34) | 82% (44) | 77% (78) | 1% (1) |
| 19. Do you have uncomfortable feelings in your stomach? | 70% (32) | 67% (35) | 66% (67) | 3% (3) |
| 20. Are you easily tired? | 65% (30) | 72% (38) | 67% (68) | 2% (2) |
| 21. Do you have thoughts about others that want to harm you? | 30% (13) | 47% (25) | 38% (38) | 4% (4) |
| 22. Do you feel more important than other people? | 10% (4) | 10% (5) | 9% (9) | 8% (8) |
| 23. Do you experience unusual/strange thoughts? | 26% (11) | 46% (24) | 35% (35) | 6% (6) |
| 24. Do you sometimes hear voices that other cannot hear? | 25% (11) | 44% (24) | 35% (35) | 3% (3) |

Abbreviations: SRQ = Self Reporting Questionnaire. \*Scores are denoted as percentage (n). The percentage resembles to percentage of ‘Yes’ answers of the total of valid (‘Yes’ or ‘No’) answers in the group.

### Determinants of Mental Health

SRQ-24 scores were significantly higher for respondents who reported adverse life events (mean=15.9 ± 4.6) than respondents who did not (mean=12.2 ±5.1); t(97)=-3.61, p<0.001. Effect size was 0.76, indicating a medium to large effect. No statistically significant associations were found between SRQ-24 scores and sex, age, length of stay in the Netherlands, presence of chronic disease, psychiatric history or substance use. However, people living on the streets (mean=15.0 ± 4.4) and in night shelters (mean=14.5 ± 5.4) tended to have higher scores (although without statistical significance) than people living in other situations, like squat locations (mean=12.8 ± 6.0) and lodging with acquaintances, friends or family (mean=12.3 ± 4.6) (F(4,82) = 0.84, p=0.50). Also, people from Central Africa (mean=18.0 ± 2.71), East Asia and Pacific (mean=17.8 ± 4.9), and Europe and Central Asia (mean=16.6 ± 2.1) tended to have higher scores than people from other regions, namely Middle East and North Africa (mean=13.6 ± 5.2), Western Africa (mean=12.8 ± 5.5), Eastern Africa (mean=12.9 ±4.9), Latin America and Caribbean (mean=10.8 ± 6.6), and South East Asia (7.00, n=1), but the overall test of variance was not statistically significant (F(7,93) = 1.77, p=0.10).

In 36 of 99 (36%) retrievable medical records, adverse life events were documented by the physician or psychologist. In the psychological help-seeking group this was 32 out of 48 (67%), versus 4 out of 51 (8%) in the general group. Some respondents reported more than 1 event. In total, 49 events were listed. The most commonly reported events were physical assault (including torture), sexual assault, captivity and combat or exposure to a war zone (Table 5).

**Table 5.** Adverse life events

| **Category\*** | **n** | **Example quote\*\*** |
| --- | --- | --- |
| **Physical assault** | 13 | *“Tortured by Al-Shabaab”* |
| **Sexual assault** | 10 | *“Raped multiple times, was in prison in Libya.”* |
| **Captivity** | 8 | *“Held captive in mortal danger.”* |
| **Combat or exposure to a war-zone** | 6 | *“He thinks about history, about the war in Somalia. Wakes up scared often. Had to flee, people around him were shot dead.”* |
| **Any other very stressful event or experience** | 5 | *“Experienced how a woman had to give birth on a plastic boat at open sea.”* |
| **Sudden violent death** | 4 | *“In the past, she saw her parents get killed.”* |
| **Sudden accidental death** | 2 | *“Mother died during travels to Netherlands”* |
| **Transportation accident** | 1 | *“Fled to Libya, crossed over by boat to Italy; the boat capsized causing 20 people to perish. Patient himself was rescued from the water after 20 hours.”* |

\*Events were categorized using the ‘Life Events Checklist for Diagnostic and Statistical Manual of Mental Disorders-5. \*\*Quotes are translated texts from electronic patient records from visits from the medical and psychological consultation hours.

### Mental Health Referrals

Twenty-nine percent of all respondents was referred to a mental health service. In the general group this was 28%, versus 30% in the psychological help-seeking group. Referred respondents had significantly higher SRQ-24 scores (mean=16.1 ± 5.5) than those who were not referred (mean=12.6 ± 4.8); t(99)=-3.21, p<0.001. Effect size was 0.68, indicating a medium to large effect.

## Discussion

### Summary of Major Findings

This study examined the prevalence of mental health problems in UMs (N=101) visiting a low-threshold primary health care service provided by an NGO. Prevalence of mental health problems and the number of referrals to mental health care services were high, also in respondents that primarily presented with physical problems only. A prevalent factor that significantly influenced mental health problems was a history of adverse life events, of which physical assault, sexual assault, captivity and combat or exposure to warzone were most common. Poor living conditions were common but not significantly related to mental health outcomes in this sample.

### Findings in Relation to Other Studies

Several studies describe poor self-reported mental health in UMs.(2) In a descriptive study among hundred female UMs in The Netherlands, more than 70% reported psychological problems including anxiety and sleeplessness.(27) In a European survey study, 22.7% of the UMs in Amsterdam reported a bad to very bad mental health, compared to 4.9% of the general population.(5) However, prevalence estimates of mental health problems in UMs differ considerably. Two retrospective studies in the Netherlands found that fewer mental health problems were registered in medical records from UMs compared to those from migrants with a residence permit. (28,29) Also, levels of psychiatric disorders amongst UMs were not elevated compared to the general population.(28,29)

The high prevalence of mental health problems observed in this study may be explained by several factors. First, the current sample differs from studies conducted in general practices. It is known that many UMs are not registered at a general practice due to several barriers in access to health care.(7,9,30,31). In addition, a qualitative exploration study showed that general practitioners tend to underreport mental health problems of UMs in their medical files.(32) Second, while UMs themselves don’t tend to easily report existing mental health issues,(7) health workers at Doctors of the World may be more focused on identifying these. Third, it is likely that the group of UMs visiting Doctors of the World included a larger proportion of rejected asylum seekers compared with the total group of UMs in the Netherlands. An earlier report showed that 67% of the patients of Doctors of the World consists of rejected asylum seekers, which is considerably higher than the most recent (but dated) estimation of 11-33% of UMs in the Netherlands.(31,33) It is plausible that rejected asylum seekers are more vulnerable for mental health problems than other UMs, like people overstaying work permits or visas. A meta-analysis in 2009 showed that prevalence estimates of major depressive disorder and anxiety disorder were almost twice as high among refugees as among labour migrants.(34) On the other hand, the stigma associated with psychological difficulties might have led to an underestimation of the prevalence of mental health problems in our study due to underreporting.(7,8,16,35)

We found a high number of adverse life events in our study. This is even more remarkable as respondents were not systematically asked about life events. In addition, consultations took place by rotating medical staff at a hardly convenient facility, which possibly contributed to underreporting. However, retrospective reporting can also lead to overreporting due to demoralization and due to active searching for explanations for the current state of mind. Despite this potential imprecision, this study found high numbers of life-events which had a medium to large effect on symptoms of common mental health disorders. Several other studies have emphasized the vulnerability of UMs and refugees to physical and sexual assault.(5,27,36) Counterintuitively and contrasting with the subjective experience of UMs themselves as found in other studies,(7,8) we found no significant correlation between poor living conditions and mental health. This may be related to the relatively small sample size in our study.

### Strengths & Limitations

A strength of this study is the unique access to this vulnerable population. The sample comprised a diverse group of UMs with regards to living situation (including night shelters, squats, living on the streets) and country of origin. Second, the study used a reliable and widely used questionnaire to assess mental health, which screens for (subclinical) mental health problems instead of using higher threshold criteria which are often used for diagnostic framing. This more adequately captures the full range of severity of mental health problems. Third, a thorough electronic patient record search was conducted, including an assessment of adverse life events.

However, certain limitations need to be addressed. This study has a limited statistical power due to a relatively small sample size and therefore small subgroups, which limited our ability to explore the association of living situations and country of origin with mental health problems. The convenience sampling method might limit generalizability, which is illustrated by the fact that regions of origin are distributed differently than in the most recent estimation report about UMs. Our study had, for example, more people from Sub-Saharan Africa, and fewer people from Asia and South-America.(1) However, due to the hidden nature of the group and the lack of available data about the population it is virtually impossible to conduct random sampling. Several factors might have led to an imprecision of results. The SRQ is not validated for a comparable population, i.e., an ethnically diverse group of UMs in a Western country. Validation studies in different populations have led to broadly varying cut-off values, ranging from ≥3 in Ethiopia to ≥17 for women in Afghanistan.(19,23) We tried to address this limitation by using the most used cut-off value of ≥8 and a more cautious value of ≥13.(21) Some questions of the SRQ did not fit our population, reflected by high numbers of missing answers for two questions, namely: ‘Is your daily work suffering?’ (29% missing) and ‘Are you unable to play a useful part in life?’ (10% missing). This is probably related to the high unemployment rate in UMs.(5) Last, due to the retrospective assessment of the electronic patient records, the number of adverse life events is probably an underestimation.

### Suggestions for Future Work

For future research, we recommend a larger study in which possible determinants for mental health are systematically investigated. Furthermore, we suggest to not just assess factors that might influence mental health negatively but also examine factors that could contribute to wellbeing. Moreover, we advise to conduct research into facilitators and barriers for psychological help in this population, to improve access to mental health care.

## New Contribution to the Literature

This study had unique access to UMs in the Netherlands. It is one of the first to quantify the mental health burden in this vulnerable group. This study underlines that mental health problems are widespread among UMs. Adverse life events were a clear influencing factor on mental health status. Experiences of physical assault like torture, sexual assault and being held in captivity were highly prevailing amongst our respondents. This study demonstrates the harshness of the past of UMs and of their current lives in a host country. In addition, it illustrates the urgency of access to health care and the need for social and psychological support. This requires a shift and thorough action from political and public health groups in order to improve health outcomes in vulnerable UMs. Endeavours such as provided by non-governmental organisations are an important first step, but more structural and institutionalized efforts are required.

## Data Availability

The data are available upon request after contacting the authors. J. Vermeulen or K. Bolhuis

